# Altered IL-6 signalling and risk of tuberculosis disease: a meta-analysis and Mendelian randomisation study

**DOI:** 10.1101/2023.02.07.23285472

**Authors:** Fergus Hamilton, Haiko Schurz, Tom A. Yates, James J. Gilchrist, Marlo Möller, Vivek Naranbhai, Peter Ghazal, Nicholas J Timpson, International Host TB Genetics Consortium, Tom Parks, Gabriele Pollara

## Abstract

IL-6 responses are ubiquitous in *Mycobacterium tuberculosis (Mtb)* infections, but their role in determining human tuberculosis (TB) disease risk is unknown. We used single nucleotide polymorphisms (SNPs) in and near the IL-6 receptor *(IL6R)* gene, focusing on the non-synonymous variant, rs2228145, associated with reduced classical IL-6 signalling, to assess the effect of altered IL-6 activity on TB disease risk. We identified 16 genome wide association studies (GWAS) of TB disease collating 17,982 cases of TB disease and 972,389 controls across 4 continents. Meta-analyses and Mendelian randomisation analyses revealed that reduced classical IL-6 signalling was associated with lower odds of TB disease, a finding replicated using multiple, independent SNP instruments and 2 separate exposure variables. Our findings establish a causal relationship between IL-6 signalling and the outcome of *Mtb* infection, suggesting IL-6 antagonists do not increase the risk of TB disease and should be investigated as adjuncts in treatment.

## Introduction

Up to a quarter of the world’s population shows evidence of exposure to *Mycobacterium tuberculosis (Mtb)* infection. Whilst only a small proportion of these individuals develop symptomatic tuberculosis (TB) disease, this translated to 10.6 million new cases and 1.6 million deaths worldwide in 2021.^1,2^ Immune responses to *Mtb* infection can determine clinical outcomes and involve multiple cellular and cytokine components with the use of IL-6 inhibitors (e.g. tocilizumab) thought to predispose to TB disease^3,4^. Contemporary clinical guidelines (e.g. American College of Rheumatology,^5^ British Society of Rheumatology^6^) and regulators (e.g. Food and Drug Administration^7^, Medicine and Health Regulatory Authority^4^) recommend screening patients for latent TB infection prior to starting tocilizumab. However, the role of this pathway in the host response to *Mtb* infection remains uncertain.^8,9^

In mice, despite promoting early Th1 responses, IL-6 is not essential for protection against *Mtb* infection^10,11^ In humans, registries of patients receiving IL-6 antagonists have not identified an elevated risk of incident TB disease.^12^ In contrast, elevated expression of plasma IL-6 cytokine and expression of an IL-6 response transcriptional signature are associated with more severe TB disease.^13,14^ Furthermore, elevated IL-6 responses are observed in patients with TB disease compared to healthy controls both at the site of *in vivo* standardised mycobacterial antigen challenge and in *ex vivo* stimulated monocytes.^15^ This increased IL-6 activity was accompanied by greater recruitment of Th17 cells and neutrophils, cells associated with tissue pathology.^15^ Elevated IL-6 responses in pulmonary TB are also associated with post-treatment lung impairment.^16^ However, whether, exaggerated IL-6 responses in people with active TB are the cause or consequence of greater tissue damage and disease severity remains unclear.

IL-6 exerts all its biological effects through binding the IL-6 receptor (IL6R) which exists in both membrane bound and soluble forms.^17^ Genetic variants in *IL6R*, which code for IL6R, associate with altered IL-6 signalling. One particular variant, rs2228145 (also known as Asp358Ala) is well-established as the major genetic determinant of IL6R levels, explaining 20-40% of observed variance.^18,19^ The minor allele (C) has a frequency of around 10% in African populations but up to 40% in other populations, such as in Europeans.^18^ The C allele is associated with alternative splicing of IL6R and increased proteolytic cleavage by ADAM17 of the membrane-bound form of IL6R, leading to an increase in soluble IL6R. The net effect of these changes is a reduction in classical *(cis)* IL-6 signalling activity and circulating levels of C-reactive protein (CRP).^19–21^ rs2228145-C is associated with reduced odds of developing several inflammatory conditions, including rheumatoid arthritis (RA) and inflammatory bowel disease (IBD).^22^ Leveraging the fact that the C allele of rs2228145 mimics (i.e. phenocopies) pharmacologic IL-6 inhibition, Mendelian randomisation (MR) analyses predicted that tocilizumab would be an effective treatment for COVID-19^23–25^ and improve cardiometabolic biomarkers in patients with high cardiovascular risk, as subsequently demonstrated in randomised controlled trials.^26,27^

In this study, we tested the hypothesis that IL-6 signalling affects the risk of developing TB disease by using genetic variation in *IL6R* as an instrumental variable in MR analyses. The available datasets allowed us to perform this analysis in several geographically diverse human populations.

## STAR Methods

### Study design

We performed a meta-analysis of genome wide association studies (GWAS) assessing host genetic susceptibility to TB disease. From these studies, we extracted the associations between variants *cis* to (within 300kb of) *IL6R* associated with IL-6 signalling and TB disease. We used these summary statistics in two sample MR analyses to estimate the causal effect of reduced IL-6 signalling on TB disease.

### Identification of GWAS and data extraction

Our inclusion criteria included all publicly available GWAS for which TB disease was an outcome, and that included the rs2228145 SNP (or SNPs in close linkage disequilibrium, R^2^ > 0.9), accepting the definition of TB used in the primary study (Table 1). We included both case-control and cohort studies, the largest of which was a multi-ancestry, GWAS meta-analysis which included 12 GWAS across diverse populations.^28^ We extracted associations from both the full meta-analysis, which was performed using MR-MEGA and GWAMA^29,30^ and each individual study. Additionally, we screened the European Bioinformatics Institute (EBI) and MRC-IEU GWAS catalogues for all GWAS on TB to identify others with publicly available summary statistics. We also searched the MEDLINE database using the terms “tuberculosis” AND “GWAS” OR “Genome wide association study”. Finally, for all included studies, we reviewed both the papers they cited and, using Google Scholar, the papers that cited them. Alongside the summary statistics, data were extracted on demographics, TB disease prevalence, TB case definitions, and methodology. Where multiple GWAS on the same population were available (e.g. UK Biobank), the most recent was used.

**Table 1.** Study populations included, TB case definitions, estimated TB prevalence and estimated TB exposure in controls. Adapted from Schurz et al. TB prevalence and estimated exposure could not be derived for additional registries and studies not part of International TB Host genetics Consortium (N/A = not available).

| Study / Dataset | TB case definition | Population | Cases / Controls | TB prevalence / 100 000 p.a. | Estimated proportion of controls ever exposed to Mtb (±SD) |
| --- | --- | --- | --- | --- | --- |
| **Studies from International TB Host genetics Consortium** |  |  |  |  |  |
| China 1 (unpublished) | Acid-fast staining and culturing of Mtb from sputum samples | Asian | 483 / 587 | 89 | 0.302 (0.101) |
| China 2 (unpublished) | Acid-fast staining and culturing of Mtb from sputum samples | Asian | 1290 / 1145 | 89 | 0.302 (0.101) |
| China 3 Qi et al, 2017 | Acid-fast staining and culturing of Mtb from sputum samples | Asian | 972 / 1537 | 89 | 0.302 (0.101) |
| Thailand - Mahasirimongkol et al 2012 | Acid-fast staining and culturing of Mtb from sputum samples | Asian | 433 / 295 | 236 | 0.404 (0.112) |
| Japan Mahasirimongkol et al 2012 | Acid-fast staining and culturing of Mtb from sputum samples. | Asian | 751 / 3199 | 23 | 0.142 (0.125) |
| Russia Curtis et al 2015 | Acid-fast staining and culturing of Mtb from sputum samples. | European | 5914 / 6022 | 109 | 0.191 (0.093) |
| Estonia (unpublished) | Biobank TB cases | European | 62 / 2660 | 13 | 0.116 (0.093) |
| Germany (unpublished) | Not available | European | 586 / 333 | 7.8 | 0.067 (0.081) |
| Gambia WTCCC | Not available | African | 1316 / 1382 | 126 | 0.280 (0.089) |
| Ghana Thye et al | Acid-fast staining and culturing of Mtb from sputum samples | African | 1359 / 1952 | 282 | 0.539 (0.198) |
| SAC(A)^ Daya et al | Acid-fast staining and culturing of Mtb from sputum samples | African | 642 / 91 | 717 | 0.436 (0.127) |
| SAC(M)* Schurz et al | Acid-fast staining and culturing of Mtb from sputum samples | African | 410 / 405 | 717 | 0.436 (0.127) |
| **Additional data sources not included in International TB Host genetics consortium** |  |  |  |  |  |
| UK Biobank | Self-report of tuberculosis | European | 2,046 / 418,427 | N/A | N/A |
| FinnGen | ICD A15 (Respiratory confirmed tuberculosis) | European | 401 / 321,302 | N/A | N/A |
| Biobank Japan | ICD A15-16 (Respiratory tuberculosis) | Asian | 549 / 211,904 | N/A | N/A |
| Zheng et al | Microbiological and clinical diagnosis | Asian | 833 / 1220 | N/A | N/A |

These searches identified 4 further relevant studies that had publicly available summary statistics: 3 cohort studies, UK Biobank (UKB)^31^, FinnGen (FG)^32^, and Biobank Japan (BBJ)^33^ and one case control study in China^34^ not included in the Schurz et al meta-analysis (Table 1). Other studies were identified but summary statistics were not available.^35–37^ Additionally, one GWAS was performed in HIV seropositive individuals, with no summary statistics available, one GWAS was performed on early progression to TB disease and so was excluded, and one GWAS merged both latent and active tuberculosis individuals, and so was also excluded.^38–40^ For UKB, TB was defined via self-report of diagnosis, and multiple GWAS have been performed by multiple groups. We chose the most recent GWAS performed by the Pan-UK Biobank team in European ancestry participants as this was performed using SAIGE^41^, which accounts for extreme case control imbalance well. In FinnGen, TB was defined using ICD-10 coding from Electronic Health Record data. Three definitions of TB were extracted – any form of TB (ICD codes A15-19), pulmonary TB (A15 or A16), and pulmonary TB with bacteriological or histological confirmation (A15 only). As we were aiming for specificity, we utilised the most specific code (A15) for downstream meta-analysis. In Biobank Japan, pulmonary TB was identified using ICD coding (ICD A15 or A16) as this was the only code available. For the study performed in China^34^, TB cases were defined using clinical and microbiological definitions (Table 1).

### Extraction of SNPs and definition of exposure

For all GWAS, we extracted effect estimates, standard errors, and allele frequencies for the rs2228145 SNP.^42^ Additionally, we extracted effect estimates for SNPs within 300kb of *IL6R*, for use in secondary analyses, described below.

### Primary analysis: CRP as instrument variable

For all studies, we performed a meta-analysis of the effect of rs2228145 on the odds of TB disease. As we only had access to GWAS summary statistics, an additive model was used, and effects were calculated per each additional C allele. Meta-analysis was performed using inverse-variance weighting (IVW) using the *meta* package in R.^43^ Heterogeneity was assessed using I^2^. Fixed effects models are reported throughout the text, although we also report random effects models in the main analysis for comparison.

MR was performed using the rs2228145 SNP as an instrument to assess the effect of reduced classical IL-6 signalling on the risk of TB disease. Our exposure was defined as reduced IL-6 signalling, as measured by the effect of this SNP on plasma CRP levels. We used estimates derived from a meta-analysis of GWAS performed in individuals from the Cohorts for Heart and Aging Research in Genomic Epidemiology (CHARGE) Consortium and UK Biobank.^21,44^ CRP as a proxy for classical IL-6 signalling has been widely used in MR studies at this locus as a phenocopy for IL-6 receptor inhibition, as CRP is produced in hepatocytes in direct response to IL-6 signalling, and variants *cis* to *IL6R* are highly likely to affect CRP by altering IL6R activity/and or function.^8,21,45,46^ The rs2228145-C allele stoichiometrically increases soluble IL6R relative to its membrane bound at both the baseline level form through differential splicing, as well as by increased efficiency of ADAM17 proteolytic cleavage of the membrane bound form in inflammation. This reduced classical signalling can potentially increase “trans” IL-6 signalling,^19–21^ and therefore this allele changes the set point of IL6-signalling but does not strictly downregulate the IL-6 pathway. However, the similarity with pharmacologic IL-6 antagonism in certain inflammatory disease contexts supports the notion that this SNP contributes towards moderated IL-6 pathway activity.^17,19,26^

MR estimates for rs2228145 were generated by the Wald ratio, and were meta-analysed using an IVW approach, as above.^47^ In line with other recent studies, we present results weighted on CRP reductions, so odds ratios (ORs) of less than one represent reduced odds of TB disease with less IL-6 signalling.

### Additional analyses: IL6R protein as instrument variable

For all studies performed on populations of European and African ancestry, we were able to perform additional analyses, using multiple SNPs cis to *IL6R* as an instrument for IL6R plasma protein levels to increase statistical power. Plasma protein levels of IL6R inversely correlate with CRP, reflecting a decrease in classical signalling, and have been used to instrument IL-6 signalling in MR studies before^48–50^. Additionally, matched instruments were available for both European and African ancestry participants, using a recent GWAS from the ARIC consortium.^18^ This GWAS (n ∼ 1,500), performed in African Americans and European Americans (independently), remains the only available GWAS data on plasma protein levels in African ancestry participants.

Independent (r^2^ <0.1) SNPs with p < 5 × 10^−8^ within 300kb of *IL6R* were extracted from both GWAS of IL6R levels performed in ARIC and used as instruments in MR independently for European and African ancestry cohorts. To ensure these SNPs had an effect on IL-6 signalling, we identified their association with high sensitivity CRP from a large recent meta-analysis^44^, and only included SNPs that had an opposing effect on CRP and IL6R (e.g. SNPs that increased IL6R and decreased CRP, or vice versa). MR estimates for each SNP were combined using IVW as our primary analytic method. For studies in Asia, we did not include any SNPs outside rs2228145 as there are no large scale GWAS of IL6R plasma protein to identify instruments. In these studies, we weighted rs2228145 by the effect on IL6R plasma protein generated in Europe, as there is little evidence of heterogeneity of effect at this exact SNP (beta in Europe 1.04, SE = 0.01); beta in Africa 1.21, SE = 0.04 per C allele), and prior data has shown similar effects of this SNP on IL-6 signalling regardless of ancestry.^50^ As sensitivity analyses, we performed other meta-analytic approaches (e.g. weighted median, MR Egger) and performed leave-one out analyses, focusing on the removal of rs2228145, to determine whether results were robust to removal of this SNP.

### Assumptions of MR

MR has 3 cardinal assumptions: relevance (that the instrument is associated with the exposure), independence (that there are no unmeasured confounders of the instrument and outcome association), and exclusion restriction (that the instrument only affects the outcome through the exposure). The relevance assumption is met in our analysis. The other two assumptions are unfalsifiable, although our sensitivity analyses (above) interrogate some of these assumptions.

The potential for confounding and horizontal pleiotropy will vary according to the outcome of interest, but these variants have been used successfully in MR studies of other infectious and inflammatory conditions, with MR results mirroring the results of subsequent RCTs.^19,26,46,51^

### Data availability

Summary data are available at the author’s GitHub (github.com/gushamilton/tb_il6r), allowing independent replication of all analyses reported.

## Results

### Study identification

From the meta-analysis by the International TB Host Genetics Consortium, we identified 12 GWAS that included rs2228145.^28^ Details of these studies have already been described but are summarised in Table 1. All were case-control studies. Four were performed in Africa,^28,52–54^ 5 in Asia,^55,56^ and 3 in Europe.^57^ Case definitions used in each study can be found in the original manuscript,^28^ but this included microbiological confirmation for most studies (9/12), except in Estonia, which was a Biobank study.

We identified 4 further relevant studies that had publicly available summary statistics: 3 cohort studies (UK Biobank (UKB)^31^, FinnGen (FG)^32^, and Biobank Japan (BBJ)^33^) and one case control study in China^34^ not included in the Schurz et al meta-analysis (Table 1). The three Biobank studies used ICD coding, while the Chinese study used microbiological confirmation to define cases of TB disease.

### Association between IL6R rs2228145-C allele and TB disease

We performed a meta-analysis across the 16 available studies, which included 18,165 cases of tuberculosis and 976,727 controls. The C allele of rs2228145 was associated with decreased risk of tuberculosis disease, with a summary odds ratio (OR) of 0.94, 95% CI 0.91 – 0.97, p = 2.92 × 10^−5^, per each additional C allele (Table 2). There was weak evidence of heterogeneity across studies (I^2^ = 0.315, p = 0.11). Effect estimates were similar in random effects analyses, although precision was reduced (OR 0.95; 95% CI 0.91 – 0.98, p = 0.009).

**Table 2:** Associations of the rs2228145-C SNP in each GWAS and meta-analysed results

| **Study** | **Odds ratio** | **Standard Error** | **C allele frequency** | **P value** |
| --- | --- | --- | --- | --- |
| **South Africa, Daya et al** | 0.991 | 0.2342 | 0.2217 | 0.9692 |
| South Africa, Schurz et al | 0.9184 | 0.147 | 0.1587 | 0.5626 |
| China 1 (unpublished) | 0.9289 | 0.1306 | 0.4247 | 0.5722 |
| China 2 (unpublished) | 0.9592 | 0.0599 | 0.4205 | 0.4868 |
| China, Qi et al | 0.8641 | 0.0606 | 0.4135 | 0.016 |
| Estonia (unpublished) | 1.1707 | 0.098 | 0.3256 | 0.9372 |
| Gambia (WTCCC) | 1.2302 | 0.1032 | 0.0905 | 0.0447 |
| Germany (unpublished) | 1.0051 | 0.1041 | 0.383 | 0.961 |
| Ghana, Thye et al | 1.0817 | 0.0876 | 0.0923 | 0.3698 |
| Japan, Mahasirimongkol et al | 0.9194 | 0.062 | 0.603 | 0.1729 |
| Russia, Curtis et al | 0.907 | 0.0287 | 0.3253 | 0.0007 |
| Finland, FinnGen | 0.8718 | 0.0875 | 0.298 | 0.117 |
| Japan, BBJ | 0.9067 | 0.0617 | 0.3928 | 0.1125 |
| China, Zheng et al | 1.011 | 0.069 | NA | 0.87 |
| UK, UKBiobank | 0.9173 | 0.0319 | 0.39 | 0.0069 |
| Thailand, Mahasirimongkol et al | 0.9996 | 0.0439 | 0.7652 | 0.9929 |
| **Meta-analysis (FE)** | **0.9405** | **0.0147** | - | **2.92 x 10<sup>-05</sup>** |
| **Meta-analysis (RE)** | **0.950** | **0.0194** | - | **0.0088** |

Next we performed MR using the rs2228145 SNP to quantify the effect of altered IL-6 signalling activity on the risk of TB disease, proxied by CRP levels. MR analyses were undertaken separately for each of the TB GWAS then pooled using fixed effects meta-analysis, yielding an OR of 0.50 (95% CI 0.39 - 0.71, p = 2.9 × 10^−5^) for each natural log CRP decrease, indicating that reduced classical IL-6 activity was associated with lower risk of TB disease development (Fig 1 & Table S1)

**Figure 1:**
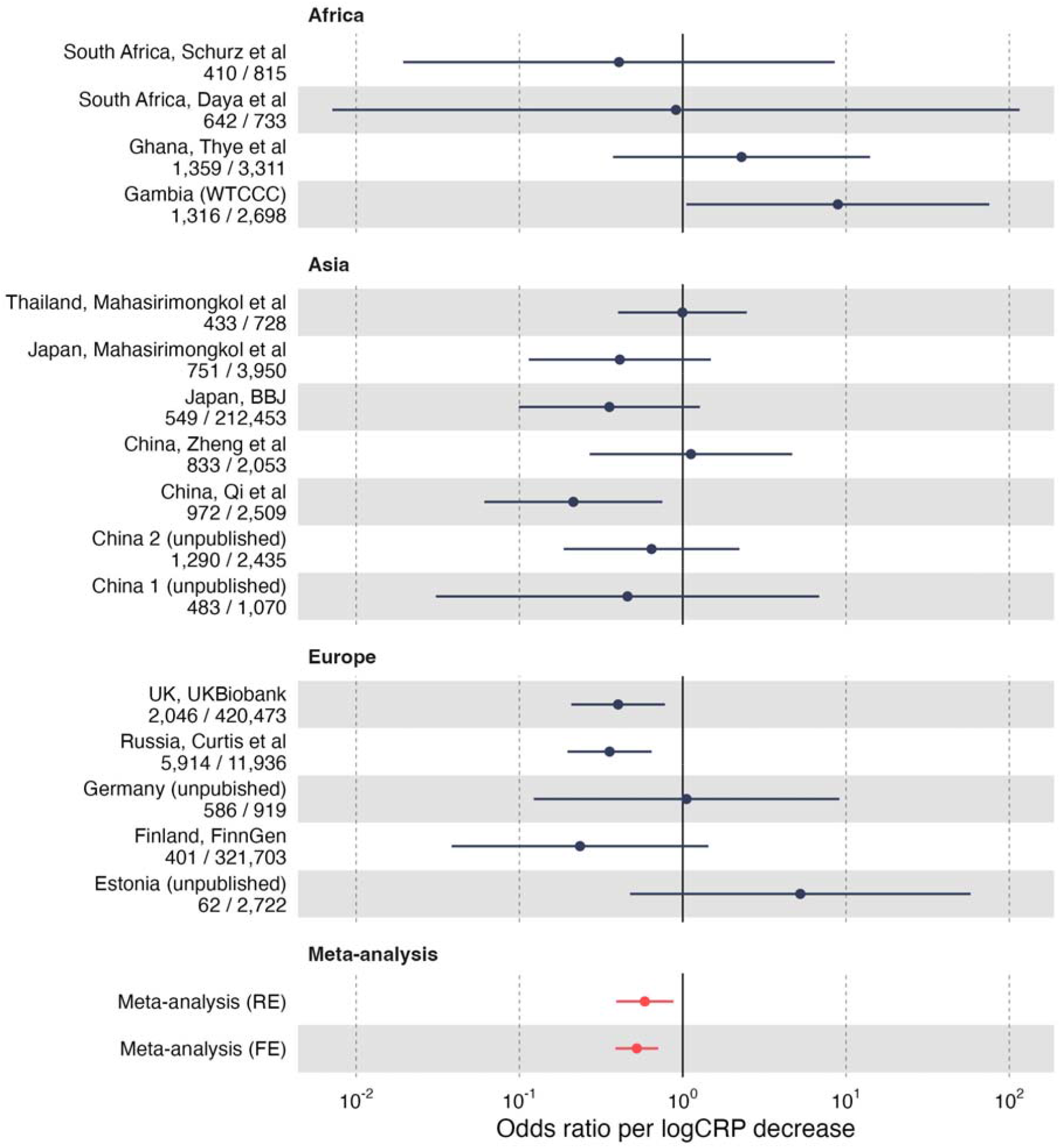
Effect of altered IL-6 signalling activty on TB disease risk using Mendelian randomisation analyses. Odds ratios for the effect of reduced IL-6 signalling (on the scale of log CRP decrease) on TB disease Estimates generated by the Wald ratio and use rs2228145 as a single instrumental variable. Odds ratios less than one suggest lower risk of TB disease.

### Association between other IL6R SNPs and TB disease

To explore the association between TB disease risk and IL-6 signalling outside rs2228145, we used alternative instrumental variables. Although rs2228145 is known to affect CRP in African ancestry populations^42,50^ there are no large scale GWAS of CRP in African ancestry populations, meaning no other SNPs can be used as instrumental variables for CRP. Therefore, we chose to use plasma concentration of IL6R protein itself as an exposure, as has been done in other MR studies at this locus.^48^ We identified independent (r^2^ <0.1) SNPs (5 in African Americans (AA), and 34 in European Americans (EA)) within 300kb of *IL6R* associated with IL6R plasma protein levels in the Atherosclerosis Risk in Communities GWAS (Table S2).^18^ To ensure these SNPs affected downstream IL-6 signalling, proxied by altered CRP concentrations, we plotted the effect of each SNP on both IL6R protein and CRP, from a large GWAS meta-analysis.^44^ As expected, there was a strong negative correlation between CRP and IL6R protein (r = −0.80 in EA, R −0.97 in AA, figure S1A&B). Only 5 SNPs were directionally inconsistent (all in European populations), and these were removed, leaving 29 to take forward to MR analyses.

Using these 29 SNPs^18^, we undertook MR analyses describing the association between plasma IL6R levels and TB disease. These analyses used data from 9 TB GWAS, all from Europe or Africa. We were unable to undertake these analyses in data from Asian GWAS, as there are no large scale GWAS of IL6R plasma protein levels available in Asian populations. These estimates using multiple SNP instruments were highly concordant with those using the single SNP rs2228145 (figure S1C), with greater precision achieved using 29 SNPs than when using rs2228145 alone.

Next, we calculated the effect of IL6R plasma protein levels on TB disease, with effect estimates from these 9 studies being derived from multiple SNPs and the remaining 7 studies from Asia (Figure 1) providing effect estimates from rs2228145 alone. As we do not have specific GWAS of IL6R in Asia, we used the effect estimates from European ancestries here, as prior data suggests little heterogeneity in effect estimates on IL-6 signalling at this specific SNP.^50^ We subsequently meta-analysed all studies by fixed effects inverse variance weighting meta-analysis. This indicated that increased levels of IL6R plasma protein, a proxy for reduced classical IL-6 signalling^19^, are associated with reduced odds of TB disease (OR 0.95; 95% CI 0.93-0.97, p = 4.15 × 10^−6^). Using rs2228145 alone yielded concordant estimates (OR 0.96; 95% CI 0.91-0.97, p = 2.92 × 10^−5^), although the use of multiple SNPs increased statistical power exemplified by a smaller p value. Full results for both models are reported in Table S3.

### Sensitivity analyses

For the 9 studies where multiple SNPs were available, we performed sensitivity analyses of the effect of IL6R plasma protein levels on odds of TB disease using different meta-analytic approaches (MR-Egger, Weighted median), which generated comparable results to those from our primary analyses using IVW (Table S4). Finally, we repeated our MR analyses describing the effect of IL6R plasma protein levels on odds of TB disease both with and without rs2228145, as this is by far the most powerful SNP. This showed comparable MR estimates from each study, although estimates lacking rs2228145 SNP were more imprecise (Figure S2).

## Discussion

To determine the role of the IL-6 pathway in human *Mtb* infection, we performed a meta-analysis of the association between genetic variation at the *IL6R* locus and odds of TB disease in 18,165 cases of tuberculosis and 976,727 controls across 16 GWAS in African, Asian and European populations. We found the C allele of the *IL6R* SNP rs2228145, which mimics the effect of IL-6 antagonists such as tocilizumab, to be associated with lower odds of TB disease. MR using both this single and other neighbouring *IL6R* SNPs as instruments supports an overarching conclusion that reduced classical IL-6 signalling is causally associated with lower risk of active TB disease.

Our study challenges current guidance indicating IL-6 inhibition confers TB disease risk. Although prospective patient registries have not identified an association between use of IL-6 antagonists and *Mtb* reactivation^3,12^, screening and treatment for latent *Mtb* infection remains recommended prior to commencement of these agents.^4–7^ The impact of such guidance is significant, including delayed initiation of biologic therapy and adverse effects of antibiotics for latent TB. Our findings indicate these concerns are unfounded, in contrast to the risk of pyogenic infections following IL-6 antagonism.^46,58^

Furthermore, our analyses indicated that lower classical IL-6 signalling may be associated with lower risk of TB disease. Strikingly, the relationship between TB and *IL6R* variants resembles that observed in the chronic inflammatory disease RA, in which the *IL6R* rs2228145 C allele is also protective, and phenocopies the therapeutic benefit of drugs inhibiting IL6R activity *in vivo*.^15^ IL-6 promotes bacterial growth and monocyte expansion following human *Mtb* infection of hematopoietic stem cells, and activity of this cytokine in blood is associated with more extensive TB disease.^13^ Tissue IL-6 activity in TB disease is associated with radiological severity and impaired lung function,^14,16^ and the activity of this cytokine can drive pathology by promoting Th17 differentiation, IL-17 activity and neutrophil chemotaxis.^17^ Indeed, this specific constellation of immune responses is elevated in TB patients compared to latent or cured TB controls following standardised *in vivo* antigen challenge.^15^ A unifying hypothesis from these observations is that exaggerated IL-6 responses associated with non-resolving *Mtb* infection increase the risk of symptomatic TB disease by promoting pathological responses, and would be consistent with the directionality of effect we observed with altered IL-6 signalling and TB disease risk.

The biology of IL-6 is complex, with both classical and trans signalling associated with disease risk.^17^ Our interpretation of the MR analyses necessitated the assumption that the rs2228145 SNP and other *cis IL6R* SNPs represent a proxy for decreased IL-6 signalling. The inverse relationship between plasma levels of CRP and IL6R is well recognised^8,50^ and all the SNPs used as instruments in this analysis associate with decreased CRP, supporting an association with an altered homeostasis set point that decreases classical IL-6 signalling. It is harder to comment on the effect of these instruments on trans signalling, given the absence of a simple biological readout for this pathway. Nevertheless, rs2228145 (and related SNPs) are still widely and successfully used as phenocopies for pharmacologic IL-6 antagonism^19,26,27^, suggesting that instrumentation of the classical signalling pathway alone allows for causal inference from these SNPs into the potential effect of IL-6 signalling on TB disease risk.

Key strengths of our study were the large and diverse patient population included alongside the robustness of the genotype association with TB disease risk using multiple cis *IL6R* SNPs. Limitations included variability in the definition of a TB case, although microbiological confirmation of *Mtb* infection alongside symptoms was a criterion for confirming TB disease cases in almost all studies included. In addition, the protective effect of the rs2228145-C allele showed some evidence of heterogeneity, with effect estimates smaller and/or reversed in some populations. Differences in TB case definitions, infecting *Mtb* strains, study design and the extent to which cases and controls are exposed to *Mtb* may all confound an apparent ancestry-specific effect. In addition, the low allele frequency of rs2228145-C in the African ancestry populations make effect estimates imprecise. Apparent heterogeneity may simply represent the play of chance, given ancestry specific heterogeneity in complex traits is rare.^59^ It is also noteworthy that MR estimates predict the effects of a lifetime of enhanced or reduced IL-6 signalling on risk of TB disease, which may qualitatively differ from the effects of short-term IL-6 antagonism ^60,61^. Although our exposure reflects a homeostasis set point change in IL-6 activity, mechanistically rs2228145-C should impact IL-6 signalling following *Mtb* exposure and MR studies using the same instrumental variable have predicted the outcome of RCTs following IL-6 blockade both in acute infection and established chronic inflammation.^5,23,27^ Finally, a TB diagnosis requires *Mtb* exposure, followed by establishment of infection that progresses to clinically apparent disease, and our data are not able to define the stage along this timeline that reduced IL-6 activity may protect from TB disease.

## Conclusions

We found genetic evidence for a causal, protective effect of reduced IL-6 signalling in the development of TB disease, observations that should diminish concerns that IL-6 blockade enhances TB disease risk. Dissecting out the precise role of IL-6 in this complex host-pathogen interaction, including the impact of TB epidemiology and host genetic background, and a possible role for IL-6 mediating pathology will require mechanistic studies involving both *in vitro* cellular models and *in vivo* human experimental medicine approaches to identify any therapeutic role for adjunctive IL-6 antagonism in human TB disease.

## Data Availability

All data produced in the present study are available upon reasonable request to the authors.

## Acknowledgments

FH’s time was funded by the GW4 CAT Doctoral Fellowship scheme (Wellcome Trust, 222894/Z/21/Z). PG’s time was funded by the Welsh Government and the EU-ERDF (Ser Cymru Scheme This work was also supported by funding from National Institute for Health Research (NIHR) (TAY, JJG, TP). NJT is a Wellcome Trust Investigator (202802/Z/16/Z) and works within the University of Bristol National Institute for Health Research (NIHR) Biomedical Research Centre (BRC). NJT is supported by the Cancer Research UK (CRUK) Integrative Cancer Epidemiology Programme (C18281/A29019). PJD was supported by a fellowship from the UK Medical Research Council (MR/P022081/1); this UK funded award is part of the EDCTP2 program supported by the European Union. ME was supported by an NHMRC fellowship (552496). The research was supported by the NHMRC grant 1025166. AvL and RvC are supported by the National Institute of Allergy and Infectious Diseases at NIH [R01 AI136921]. AM and RM are funded by the EU project no. 2014-2020.4.01.15-0012 “Gentransmed”. BA is supported by the ‘Scientific Programme Indonesia Netherlands’ (SPIN) under the Royal Academy of Arts and Sciences (KNAW), the Netherlands. TP was funded by the Wellcome Trust 222098/Z/20/Z). GP’s time was funded by the UCLH NIHR Biomedical Research Centre. The views expressed in this publication are those of the author(s) and not necessarily those of the NHS, the National Institute for Health Research or the Department of Health and Social Care.

## Author contributions

Study conception, design, and oversight, FH, TP and GP; data acquisition, FH, HS, TAY, JJG, MM, VN, TP; data analysis and interpretation, FH, HS, TP, GP; manuscript composition and drafting FH, TAY, TP and GP. All authors reviewed and approved the final version of this manuscript.

## Ethics statement

This study makes use of publicly available summary genetic association data which are available from peer reviewed publications or from the preprint of the ITHGC.^28^ All institutions involved in the ITHGC have ethics approval for their respective studies: China 1 and 2: The study protocol was approved by the Ethics Committee of the Beijing Chest Hospital, the 309 Hospital of the PLA, Shijiazhuang Fifth Hospital, the China PLA General Hospital, the Tongliao TB institute and the Center for Diseases Control and Prevention in Jalainuoer. China 3: Ethics approval was granted by the Ethics Committees of the Beijing Childrens Hospital, the Beijing Geriatric Hospital, the Tuberculosis Hospital in Shaanxi Province, the Beijing Institute of Genomics, Chinese Academy of Sciences and the Center for Disease Control and Prevention of Jiangsu Province. Thailand: Ethics approval was granted by the Ethics Review Committee of the Ministry of Public Health in Thailand. Japan: Ethics approval was granted by the Institutional Review Board of the Center for Genomic Medicine, RIKEN Russia: Blood samples from all participants were collected and studied with written informed consent according to the Declaration of Helsinki and with approvals from the local ethics committees in Russia (St. Petersburg and Samara) and the UK (Human Biological Resource Ethics Committee of the University of Cambridge and the National Research Ethics Service, Cambridgeshire 1 REC, 10/H0304/71). Estonia: The Estonian Bioethics and Human Research Council (EBIN) approved the Estonian Genome Center study reported in this manuscript. Germany: The study protocol was approved by the ethics committee (EC) of the University of Luebeck, Germany (reference 07/125), and was adopted by other ethics committees covering all 18 participating centres (EC of the medical faculty of the University of Goettingen; EC of the Medical Council of Hessen, Frankfurt /Main; EC of the Medical Council Hamburg; EC of the Medical Council Lower Saxony, Hannover; EC of the Medical Faculty Carl Gustav Carus, Technical University of Dresden; EC of the Medical Council Berlin; EC of the Medical Council Bavaria, Munich; EC of the Medical Faculty, Friedrich Alexander University Erlangen Nuremberg; EC of the Medical Faculty of the University of Regensburg; EC of the University of Witten/ Herdecke) Gambia: Ethics approval was granted by the Medical Research Council (MRC) and the Gambian government joint ethical committee. Ghana: Ethics approval was granted by the Committee on Human Research, Publications and Ethics, School of Medical Sciences, Kwame Nkrumah University of Science and Technology, Kumasi, Ghana, and the Ethics Committee of the Ghana Health Service, Accra, Ghana. SAC A and SAC M: Ethics approval was granted by the Health Research Ethics Committee of Stellenbosch University (project registration numbers S17/01/013, NO6/07/132 and 95/072).

## Notes

### Competing Interest Statement

The authors have declared no competing interest.

### Funding Statement

FH time was funded by the GW4 CAT Doctoral Fellowship scheme (Wellcome Trust, 222894/Z/21/Z). PG time was funded by the Welsh Government and the EU-ERDF (Ser Cymru Scheme This work was also supported by funding from National Institute for Health Research (NIHR) (TAY, JJG, TP). NJT is a Wellcome Trust Investigator (202802/Z/16/Z) and works within the University of Bristol National Institute for Health Research (NIHR) Biomedical Research Centre (BRC). NJT is supported by the Cancer Research UK (CRUK) Integrative Cancer Epidemiology Programme (C18281/A29019). PJD was supported by a fellowship from the UK Medical Research Council (MR/P022081/1); this UK funded award is part of the EDCTP2 program supported by the European Union. ME was supported by an NHMRC fellowship (552496). The research was supported by the NHMRC grant 1025166. AvL and RvC are supported by the National Institute of Allergy and Infectious Diseases at NIH [R01 AI136921]. AM and RM are funded by the EU project no. 2014-2020.4.01.15-0012 Gentransmed. BA is supported by the Scientific Programme Indonesia Netherlands (SPIN) under the Royal Academy of Arts and Sciences (KNAW), the Netherlands. TP was funded by the Wellcome Trust 222098/Z/20/Z). GPs time was funded by the UCLH NIHR Biomedical Research Centre. The views expressed in this publication are those of the author(s) and not necessarily those of the NHS, the National Institute for Health Research or the Department of Health and Social Care.

### Author Declarations

This study makes use of publicly available summary genetic association data which are available from peer reviewed publications or from the preprint of the International Tuberculosis Host Genetics Consortium (ITHGC) - https://www.medrxiv.org/content/10.1101/2022.08.26.22279009v2. All institutions involved in the ITHGC have ethics approval for their respective studies: China 1 and 2: The study protocol was approved by the Ethics Committee of the Beijing Chest Hospital, the 309 Hospital of the PLA, Shijiazhuang Fifth Hospital, the China PLA General Hospital, the Tongliao TB institute and the Center for Diseases Control and Prevention in Jalainuoer. China 3: Ethics approval was granted by the Ethics Committees of the Beijing Childrens Hospital, the Beijing Geriatric Hospital, the Tuberculosis Hospital in Shaanxi Province, the Beijing Institute of Genomics, Chinese Academy of Sciences and the Center for Disease Control and Prevention of Jiangsu Province. Thailand: Ethics approval was granted by the Ethics Review Committee of the Ministry of Public Health in Thailand. Japan: Ethics approval was granted by the Institutional Review Board of the Center for Genomic Medicine, RIKEN Russia: Blood samples from all participants were collected and studied with written informed consent according to the Declaration of Helsinki and with approvals from the local ethics committees in Russia (St. Petersburg and Samara) and the UK (Human Biological Resource Ethics Committee of the University of Cambridge and the National Research Ethics Service, Cambridgeshire 1 REC, 10/H0304/71). Estonia: The Estonian Bioethics and Human Research Council (EBIN) approved the Estonian Genome Center study reported in this manuscript. Germany: The study protocol was approved by the ethics committee (EC) of the University of Luebeck, Germany (reference 07/125), and was adopted by other ethics committees covering all 18 participating centres (EC of the medical faculty of the University of Goettingen; EC of the Medical Council of Hessen, Frankfurt /Main; EC of the Medical Council Hamburg; EC of the Medical Council Lower Saxony, Hannover; EC of the Medical Faculty Carl Gustav Carus, Technical University of Dresden; EC of the Medical Council Berlin; EC of the Medical Council Bavaria, Munich; EC of the Medical Faculty, Friedrich Alexander University Erlangen Nuremberg; EC of the Medical Faculty of the University of Regensburg; EC of the University of Witten/ Herdecke) Gambia: Ethics approval was granted by the Medical Research Council (MRC) and the Gambian government joint ethical committee. Ghana: Ethics approval was granted by the Committee on Human Research, Publications and Ethics, School of Medical Sciences, Kwame Nkrumah University of Science and Technology, Kumasi, Ghana, and the Ethics Committee of the Ghana Health Service, Accra, Ghana. SAC A and SAC M: Ethics approval was granted by the Health Research Ethics Committee of Stellenbosch University (project registration numbers S17/01/013, NO6/07/132 and 95/072).

