## Supplementary figures for "Altered IL-6 signalling and risk of tuberculosis disease: a meta-analysis and Mendelian randomisation study"

**Figure S1:** Relationship between IL6R SNPs and TB disease risk using IL6R protein as an alternative instrument variable. Effect of IL6R SNPs on IL6R protein and CRP in populations of A) European ancestry and B) African ancestry. IL6R betas derived from ARIC GWAS, CRP betas derived from meta-analysis of GWAS performed in individuals from CHARGE and UK Biobank. C) MR-derived odds ratios for the effect of downregulated IL-6 signalling (on the scale of log IL6R plasma protein) on TB disease, with MR estimates from IVW-weighted multiple SNPs (red) compared to Wald ratio-derived estimates using only rs2228145 (black). Multiple SNP analyses in C) used 5 and 27 SNPs for African and European studies respectively.

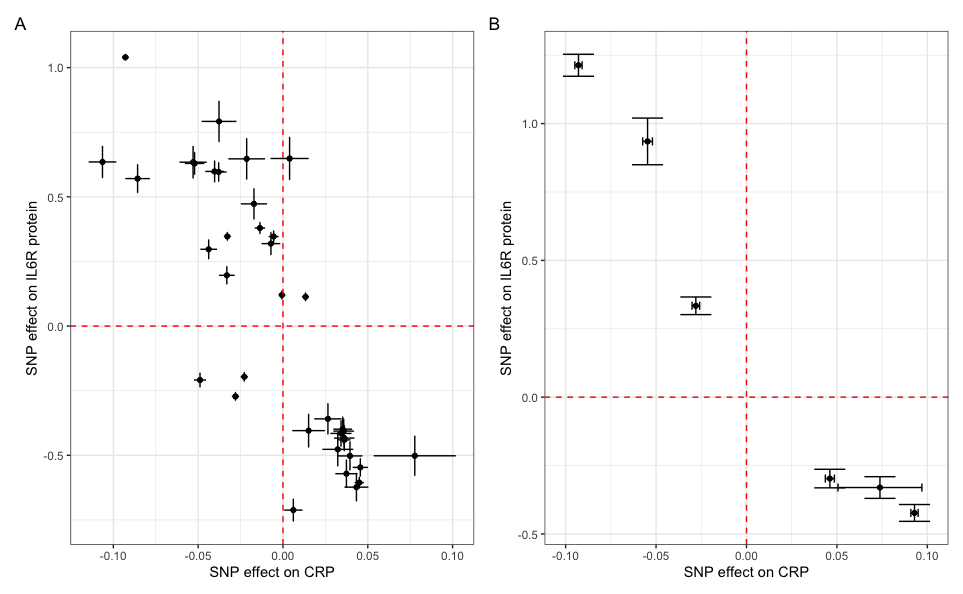

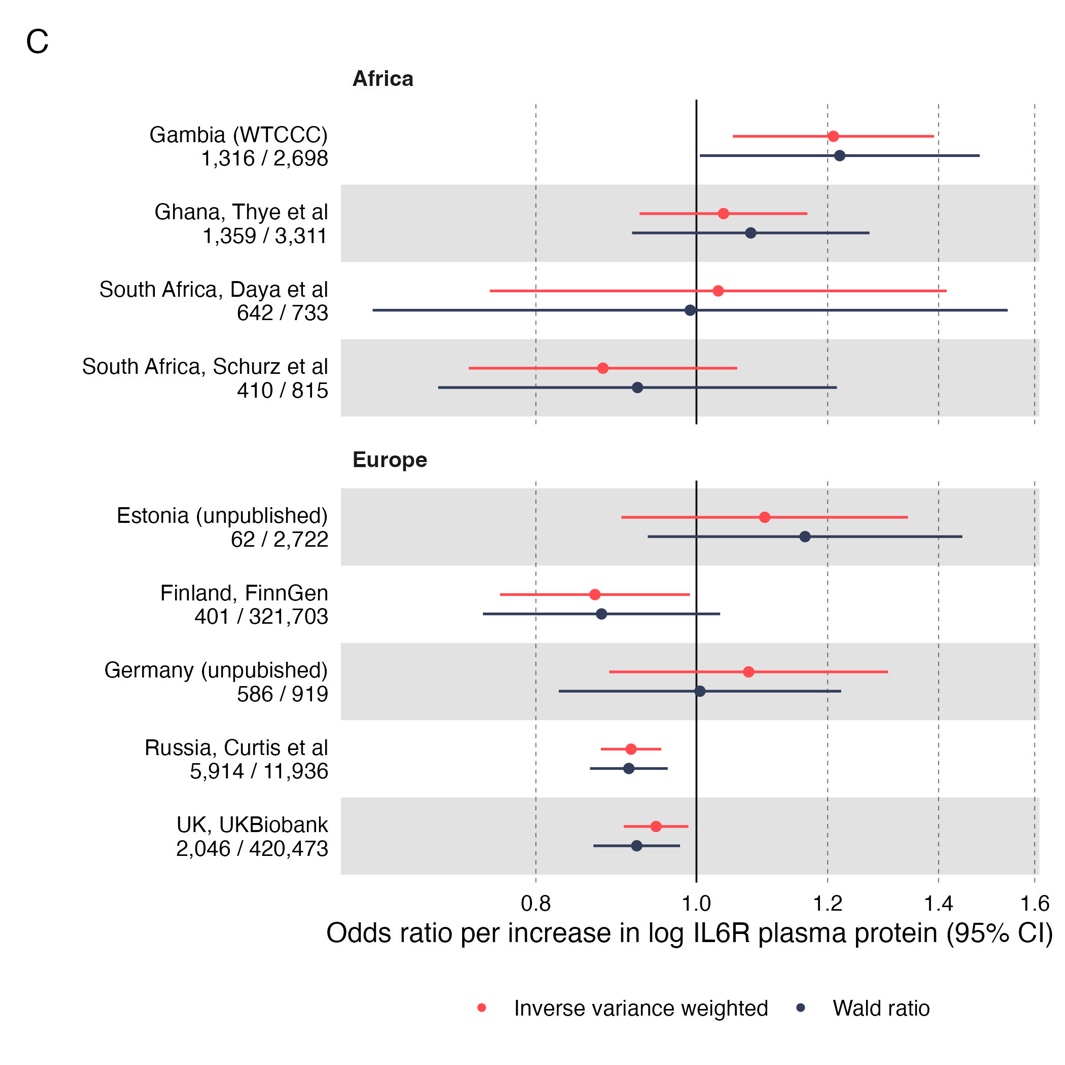

**Figure S2:** Effect of removing rs2228145 in a leave-one-out inverse variance weighting MR analysis. This was performed for all studies (9/16) where multi-ancestry cis pQTLS were available for IL6R plasma protein levels. Red represents the IVW estimate excluding rs2228145, while black is the estimate including rs2228145.

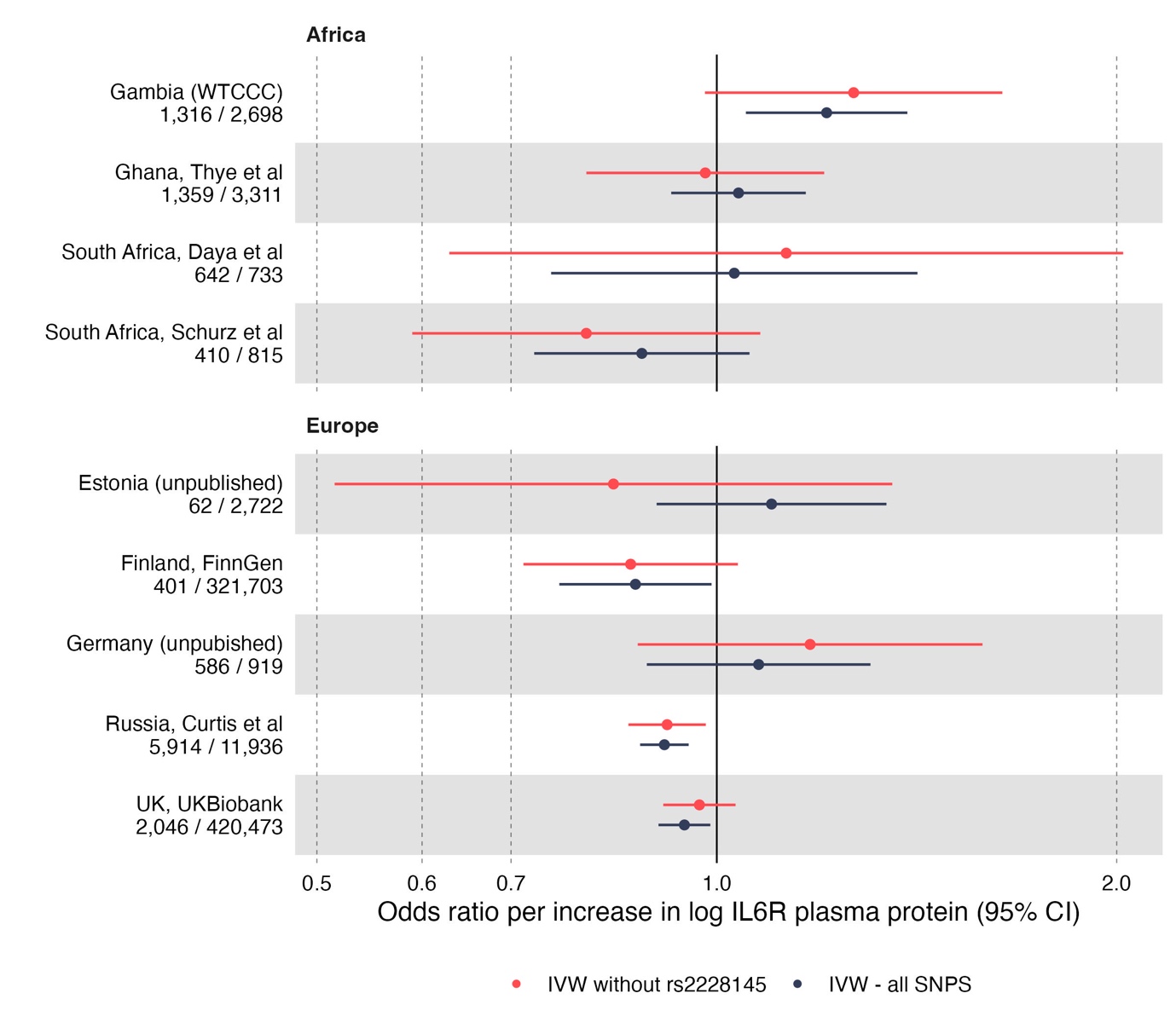
